# Gut microbiota development in very preterm infants following fortification of human milk

**DOI:** 10.1101/2024.05.07.24306981

**Authors:** Lin Yang, Yan Hui, Per Torp Sangild, Witold Piotr Kot, Lise Aunsholt, Gitte Zachariassen, Ping-Ping Jiang, Dennis Sandris Nielsen

## Abstract

**Background:** Very preterm infants (VPIs) are born with an immature gut, being sensitive to gut microbiota dysbiosis-related disease like necrotizing enterocolitis. While human milk is the best source of nutrition for VPIs, it requires fortification to meet their nutrient requirements for optimal growth. However, the optimal type of fortifier remains uncertain. Bovine colostrum (BC), rich in protein and bioactive components, may be an alternative to conventional fortifiers (CF). We aimed to investigate the distinct impacts of different bovine fortifiers, BC and CF, on the gut microbiota of VPIs. The gut microbiota of 225 VPIs who were fed human milk fortified with either BC or CF, were profiled by 16S rRNA gene amplicon sequencing of fecal samples collected before, one and two weeks of fortification.

**Results:** Fortifier type affected the microbial community structure to a modest extent, but only explaining 1% of the variance, and no specific taxa differed between the BC and CF groups. This fortifier-derived impact was predominantly observed in VPIs born via caesarean section. Birth mode exhibited transient effects on microbial community structure shortly after birth, with caesarean section-born VPIs dominated by *Firmicutes*, while vaginally-born VPIs were dominated by *Proteobacteria*. This birth mode-derived difference diminished with age and disappeared around one month after birth. The fecal pH, increased by BC, was positively correlated with *Staphylococcus* and *Corynebacterium*, and negatively with *Bifidobacterium* abundance. The change in relative abundance of *Staphylococcus* was negatively correlated with weight gain.

**Conclusion:** Collectively, fortification of human milk with BC or CF does influence the gut microbiota of VPIs but only to a modest extent during early life. Conversely, birth mode appears to be a significant temporary factor influencing the gut microbiota during this period.

Our findings are consistent with existing literature and support the idea that the choice of fortifier has limited effects on gut microbiota development in the first month of life of VPIs.

## Background

Very preterm infants (VPIs, born before 32 weeks of gestation) are characterised by having immature organs including the gut and immune systems, and are highly susceptible to extrauterine growth restriction (EUGR) and gut and immune complications, such as necrotizing enterocolitis (NEC) and late-onset sepsis (LOS) [1,2]. Mother’s own milk (MOM) and donor human milk (DHM) are the preferred sources of nutrients for VPIs, but do not contain adequate nutrients and energy for optimal growth thus requiring nutrient fortification [3]. Most nutrient fortifiers are formula products based on bovine milk (bovine milk-based fortifiers, BMBFs), generally being subject to extensive processing involving protein pre-hydrolysis and often added plant-based ingredients [4]. Unfortunately, such fortifiers have been associated with an increased risk of gut-related complications [5]. Human milk-based fortifiers (HMBFs) have been developed but are not widely available and clinical benefits are not consistent [6–9]. Bovine colostrum (BC), the milk delivered within the first day after parturition in cows, has a high level of protein and contains many bioactive factors with antibacterial and immunomodulatory activities [10]. Using preterm piglets as model for preterm infants, BC has been found to increase body growth and prevent gut complications when fed exclusively as well as when used as a supplement to human milk [11–15]. A pilot trial (n = 50) showed that BC supplemented to MOM was well tolerated in VPIs [16,17]. In two subsequent, larger randomized controlled trials (RCTs), the feasibility of using BC as a supplement to MOM in the first weeks of life (compared with preterm formula as supplement to MOM, n = 350) [18], or as a fortifier to human milk later (compared with a conventional fortifier, CF, n = 232) [19], was evaluated. Both trials showed that VPIs supplemented with BC had similar growth and clinical outcomes as VPIs fed formula and CF [18,19].

At and around birth, the gut is colonized by a myriad of microorganisms. Compared with that of term born infants, the gut microbiota (GM) of VPIs is characterised by lower species diversity [20,21], a higher abundance of facultative anaerobes like *Staphylococcus*, *Enterococcus*, *Klebsiella*, *Enterobacter* and *Escherichia* and a lower abundance of bifidobacteria and lactobacilli [22,23]. The aberrant GM in VPIs is associated with an increased risk of morbidities, such as NEC and LOS, and impaired growth and neurodevelopment [24–26]. Yet, it remains unclear whether specific GM changes are directly implicated in inducing NEC and LOS, and how this relates to nutrient maldigestion. Birth mode, gestational age (GA), hospital environment, the type of enteral nutrition (EN) and exposure to antibiotics and probiotics have all been reported to influence the GM in preterm infants [25,27–29]. Furthermore, the GM of VPIs receiving human milk (MOM and/or DHM) differs from that in infants fed preterm formula, harbouring less *Escherichia* and *Clostridium* though the effect on GM composition tends to be modest [8,30,31]. In a pilot study, BC supplementation decreased the relative abundance of *Lactobacillaceae* and *Enterococcaceae* in VPIs, relative to DHM [32].

Compared with highly processed BMBFs, a mildly heat-treated fortifier like BC, containing intact milk proteins (casein, whey and immunoglobulins), lipids, lactose and many bioactive components [10,19], may have different effects on GM development relative to CF [32]. In a recent randomized clinical trial where BC was compared with CF (containing hydrolysed whey protein) as fortifier to human milk for VPIs, both fortifiers improved growth to the desired level and morbidities were similar between the two groups of infants receiving different fortifiers, though the study was not powered to detect differences in morbidities [19]. Importantly, bowel habits tended to be improved after the entire intervention period (about 1 month) for BC infants [33]. In this sub-analysis, we aim to extend these findings to investigate whether fortification with BC or CF affects the GM in the weeks just after start of fortification where the concern for gut complications, such as feeding intolerance, GM dysbiosis, and NEC, is particularly high.

## Results

Twenty-nine and five samples were excluded due to unmatched collection time and/or low sequencing reads.

The FortiColos trial (Bovine colostrum as a fortifier added to human milk for preterm infants, clinicaltrial.gov: NCT03537365) is a multicentre, randomised clinical trial conducted at eight hospitals in Denmark testing the feasibility of using BC as nutrient fortifier to MOM and/or DHM. A total of 232 VPIs were enrolled and randomized to receive either BC or CF as a fortifier added to human milk until postmenstrual age (PMA) 34+6 weeks, starting when the enteral nutrition volume reached 100-140 mL/kg/d and blood urea nitrogen (BUN) < 5 mmol/L [19]. Fecal samples were collected before (fortification time-point 0, FT0), and one (FT1) and two weeks (FT2) after start of fortification (**Fig. 1a**). FT0, FT1 and FT2 were roughly equivalent to 2-10, 10-20 and 20-30 days of life (DOL, **Fig. S1b)**. In total, 558 fecal samples from 225 infants (BC, n = 109; CF, n = 116) with acceptable data quality after 16S ribosomal RNA (rRNA) gene amplicon sequencing were included for analysis (192, 191 and 175 samples at FT0, FT1 and FT2, respectively, **Fig. 1b**).

**Fig 1.**
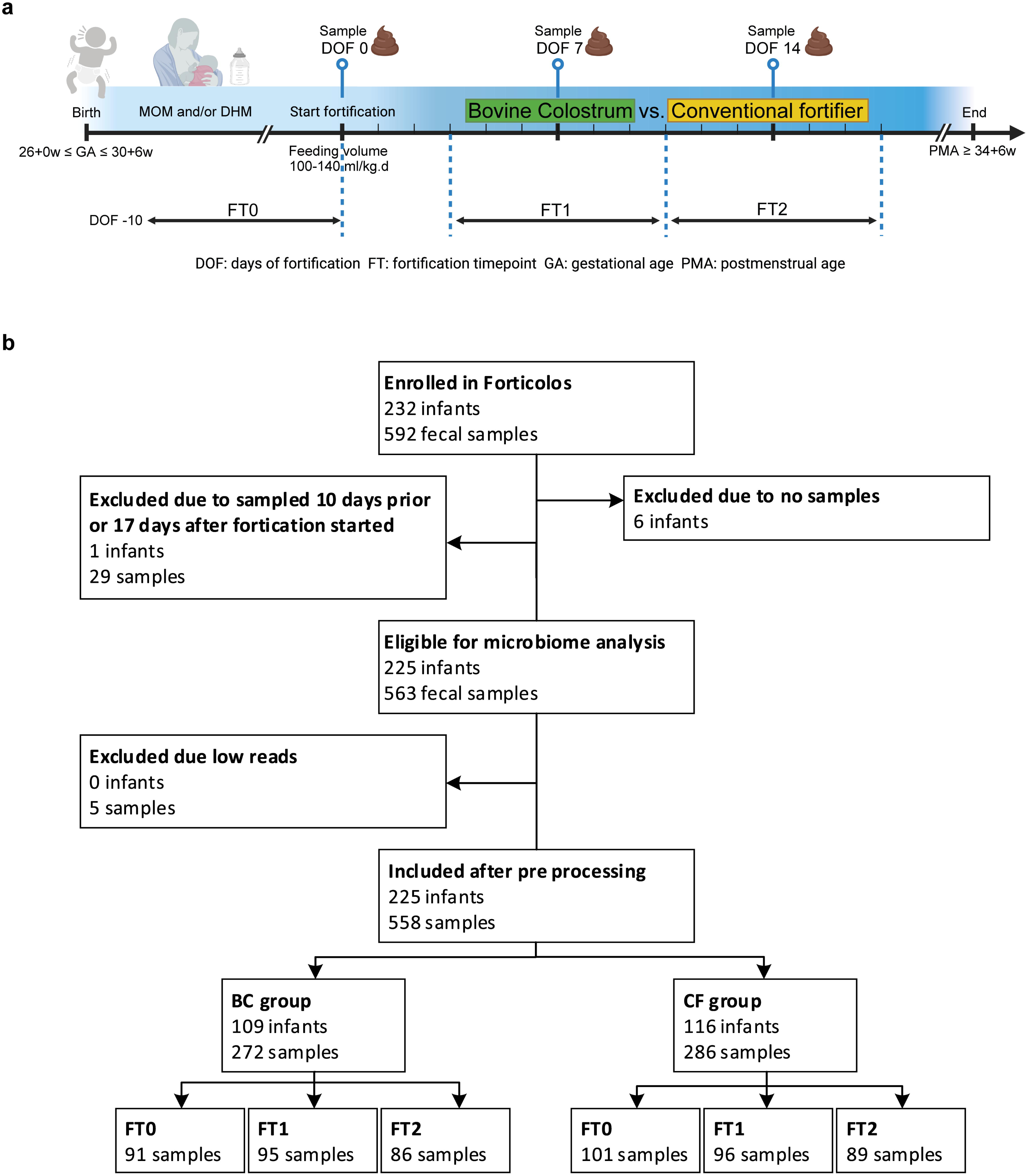
Design of the FortiColos trial and sample inclusion for the microbiota analysis. **a** Study design and the faecal sample collection. Faecal samples before fortification (from -10 to 0 days of fortification, DOF, FT0), around one week (7 ± 3 DOF, FT1) and two weeks of fortification (14 ± 3 DOF, FT3) were included. Created with BioRender.com. **b** Infants (samples) inclusion for the microbiota analysis. Twenty-nine and five samples were excluded due to unmatched collection time and/or low sequencing reads.

### Characteristics of the included infants

Characteristics of the 225 infants with eligible samples for microbiota analysis are shown in **Table 1**. No significant difference was found in GA, birth weight (BW), sex (male or female), small for gestational age (SGA, yes or no), multiple births (singleton or not), birth mode (caesarean section, CS or vaginal birth, VB) or 3-day MOM proportion (the number of meals of exclusive MOM over the number of all meals in the three days before a specific FT) between the infants receiving BC or CF (all *p* > 0.05, **Table 1**). No significant difference was found between the BC and CF groups in the numbers of infants receiving probiotics at hospitals with routine probiotics use before fortification, or in the numbers of infants receiving antibiotics at any FT (all *p* > 0.05, **Table 1**). The fraction of infants born by CS was not different between the two groups. No clear phenotypic differences were observed between infants born by CS or VB, except that the prevalence of infants SGA were higher in the CS infants (p < 0.05, **Table S1**).

**Table 1.**
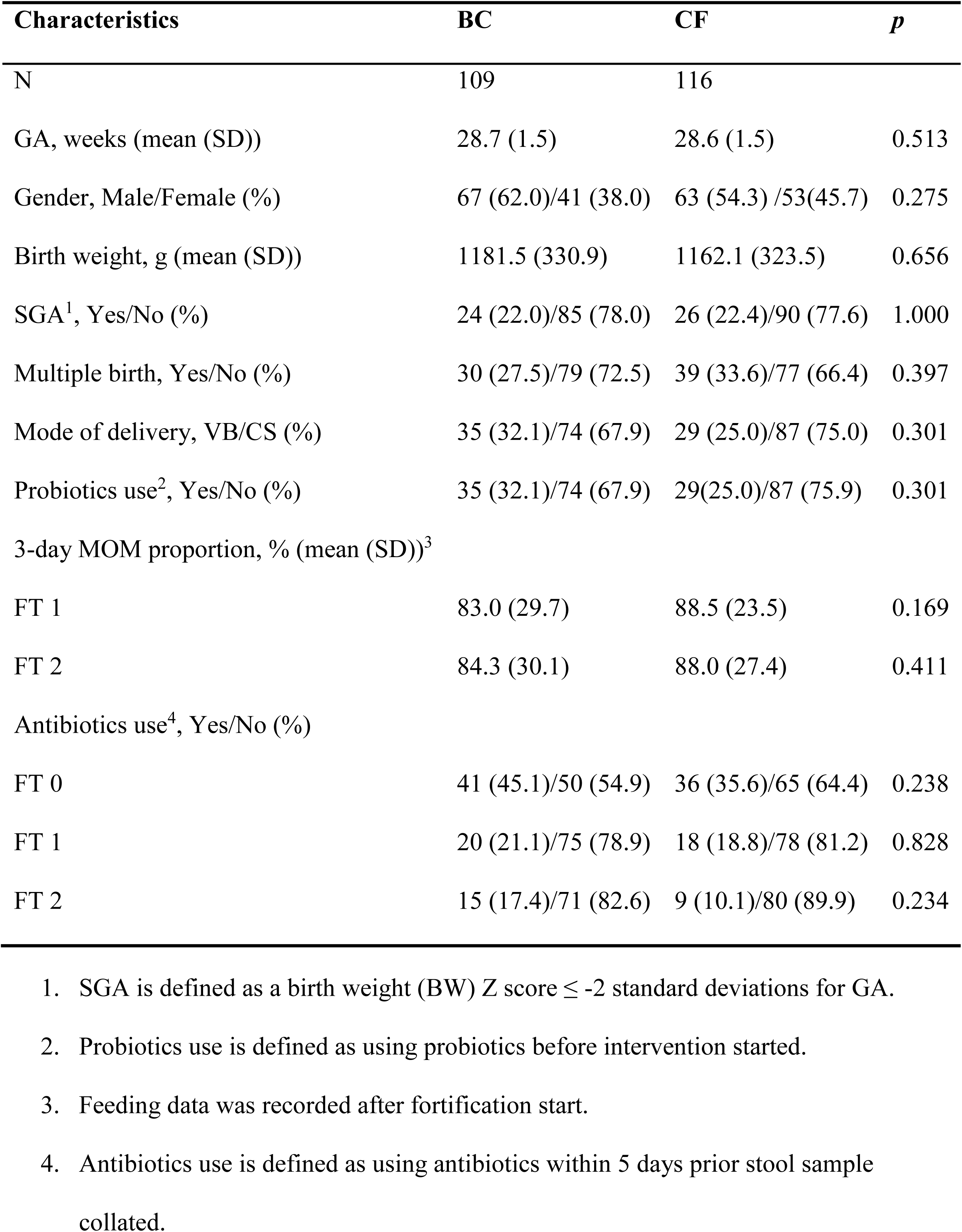
Characteristics of infants included for microbiota analysis.

| Characteristics | BC | CF | *p* |
| --- | --- | --- | --- |
| N | 109 | 116 |  |
| GA, weeks (mean (SD)) | 28.7 (1.5) | 28.6 (1.5) | 0.513 |
| Gender, Male/Female (%) | 67 (62.0)/41 (38.0) | 63 (54.3) /53(45.7) | 0.275 |
| Birth weight, g (mean (SD)) | 1181.5 (330.9) | 1162.1 (323.5) | 0.656 |
| SGA<sup>1</sup>, Yes/No (%) | 24 (22.0)/85 (78.0) | 26 (22.4)/90 (77.6) | 1.000 |
| Multiple birth, Yes/No (%) | 30 (27.5)/79 (72.5) | 39 (33.6)/77 (66.4) | 0.397 |
| Mode of delivery, VB/CS (%) | 35 (32.1)/74 (67.9) | 29 (25.0)/87 (75.0) | 0.301 |
| Probiotics use<sup>2</sup>, Yes/No (%) | 35 (32.1)/74 (67.9) | 29(25.0)/87 (75.9) | 0.301 |
| 3-day MOM proportion, % (mean (SD))<sup>3</sup> |  |  |  |
| FT 1 | 83.0 (29.7) | 88.5 (23.5) | 0.169 |
| FT 2 | 84.3 (30.1) | 88.0 (27.4) | 0.411 |
| Antibiotics use<sup>4</sup>, Yes/No (%) |  |  |  |
| FT 0 | 41 (45.1)/50 (54.9) | 36 (35.6)/65 (64.4) | 0.238 |
| FT 1 | 20 (21.1)/75 (78.9) | 18 (18.8)/78 (81.2) | 0.828 |
| FT 2 | 15 (17.4)/71 (82.6) | 9 (10.1)/80 (89.9) | 0.234 |

1. SGA is defined as a birth weight (BW) Z score $\leq -2$ standard deviations for GA. 2. Probiotics use is defined as using probiotics before intervention started. 3. Feeding data was recorded after fortification start. 4. Antibiotics use is defined as using antibiotics within 5 days prior stool sample collated.

### Birth mode affects gut microbiota composition of very preterm infants

Before assessing the effect of fortifier on VPI GM development, we investigated if other variables including fortifiers, GA, SGA, birth mode, DOL, hospital and use of antibiotics significantly influenced GM. Data is also available on administration of probiotics to the VPIs, but probiotic use is fully confounded by “hospital”, as the involved hospitals either administer probiotics to all VPIs (4 hospitals) or none at all (4 hospitals). “Hospital” was found to significantly influence VPI GM, but whether this difference is driven by the use of probiotics, other hospital specific factors or both is not possible to determine (**Fig. S2**, **Table S2**). Among all investigated variables, birth mode was the variable that had the most pronounced influence on VPI GM composition, as shown by distance-based redundancy analysis (dbRDA) (**Table S3**), prompting a more in-depth investigation.

Of the VPIs receiving BC, 35 were VB and 74 were born by CS, while among the CF fed VPIs, 29 were VB and 87 were born by CS. Birth mode (CS or VB) did not affect the species diversity as determined by the Shannon diversity index or the number of observed zero-radius Operational Taxonomic Units (zOTUs) at any FT (all *p* > 0.05, **Fig. S3a, S3b**). However, the microbial community structure differed between the CS and VB infants in the first 20 days postpartum (FT0, R^2^ = 0.05; FT1, R^2^ = 0.02, respectively, both *p* = 0.001, **Fig. 2a, 2b**) but at FT2, the effect was smaller (R^2^ = 0.01) and not significant (FT2, *p* = 0.06, **Fig. 2c**), indicating a decreasing influence of birth mode on GM as the VPI gets older over the first weeks.

**Fig 2.**
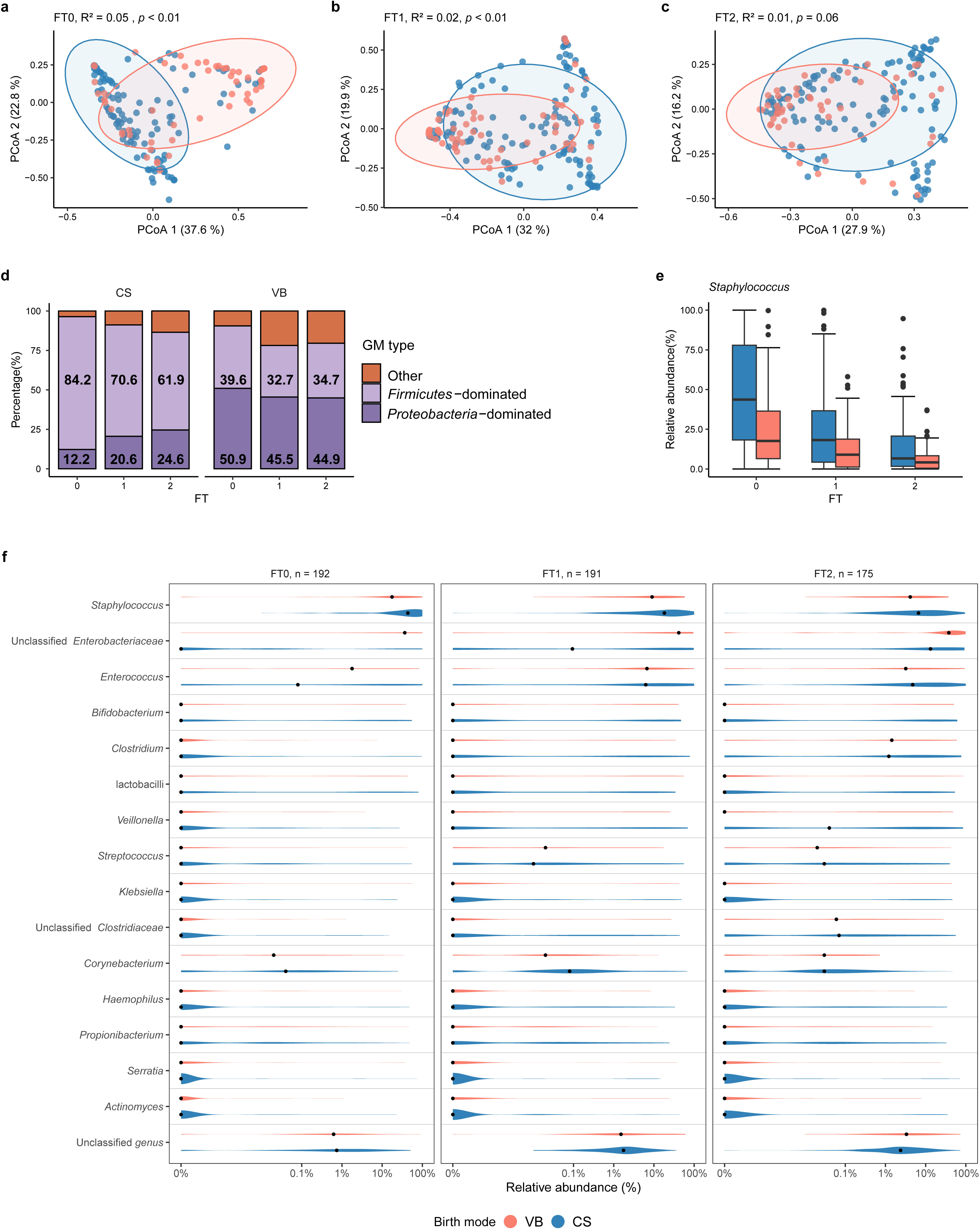
Birth mode transiently affects the gut microbiota. **a-c** The microbial community structure was different in VPIs born by caesarean section (CS) or vaginal birth (VB) but the difference diminishes over time. PCoA plots were based on weighted Unifrac dissimilarity metrics. The ellipses represented 95% confidence intervals. R^2^ and *p* values were calculated by PERMANOVA (999 permutations) and adjusted for confounders, including GA, SGA, DOL, fortification, hospital and use of antibiotics. **d** CS infants were most *Firmicutes*-dominated while VB infants were most *Proteobacteria*- dominated. The GM type of the CS infants changed over time. The GM type of the CS and VB infants at the same FT was compared by chi-square (χ^2^) test. The comparison of infants being *Firmicutes*-dominated or not between different FTs was conducted using logistic regression with adjustments for confounders within each birth mode group. **e** Relative abundance of *Staphylococcus* was different in the CS and VB infants. The relative abundance of *Staphylococcus* was higher in the CS group at FT1 and FT2, as revealed by DESeq2 with adjustment of confounders before FDR correction at each FT. **f** Relative abundance of the most abundant 15 genera in the CS and VB infants at three FTs. The order of genera was based on the mean abundance. Median values are shown as black dots. A pseudocount of 1 × 10^-6^ was added to all relative abundance values to facilitate log-scale transform.

Most CS infants had a *Firmicutes*-dominated GM (with 84.2% of infants being *Firmicutes* dominated at FT0 and 61.9% at FT2), while most VB infants had a GM dominated by *Proteobacteria* (44.9-50.9%) at all FTs (all *p* < 0.01, **Fig. 2d**). The number of *Firmicutes*- dominated infants decreased with time in the CS infants (FT0 vs FT1, FT0 vs FT2, both *p* < 0.05, **Fig. 2d**), but not in the VB infants (FT0 vs FT1 vs FT2, all *p* > 0.05, **Fig. 2d**). At the genus level, *Staphylococcus, Enterococcus* and unclassified *Enterobacteriaceae* were the three most abundant genera at all three FTs (**Fig. 2f**). The relative abundance of *Staphylococcus* was significantly higher in the CS infants than that in the VB infants at FT1 and FT2, before (both *p* < 0.05), but not after (q > 0.05, DESeq2, **Fig. 2e**) false discovery rate (FDR) correction. Interestingly, the relative abundance of *Bacteroidetes* was above 2% in VB infants, while in CS infants, this phylum was either not detected or very low abundant (<0.01%) at any FT **(Fig. S3c).**

### Choice of fortifier has limited effect on GM development during the first month of life for very preterm infants

No significant difference was found between the BC and CF groups at any FT with respect to the Shannon diversity index or the number of observed zOTUs (all *p* > 0.05, **Fig. 3d, 3e**). However, the gut microbial community structure differed between the BC and CF groups at FT1 and FT2 (both *p* = 0.01, **Fig. 3b, 3c**) as determined by Weighted UniFrac distance metrics, but choice of fortifier only explained a relatively small proportion of the variance (R^2^ = 0.01 at both FTs, **Fig. 3b, 3c**). Similar results were found when other (dis)similariy metrics, such as unweighted UniFrac, Bray-Curtis and Binary Jaccard, were used (**Fig. S4**). In a subgroup analysis by birth mode, the fortifier-driven difference in the microbial community structure was only observed in the infants born by CS (*p* < 0.05, **Fig. S5**), but the variance explained was still modest (R^2^ = 0.02).

**Fig 3.**
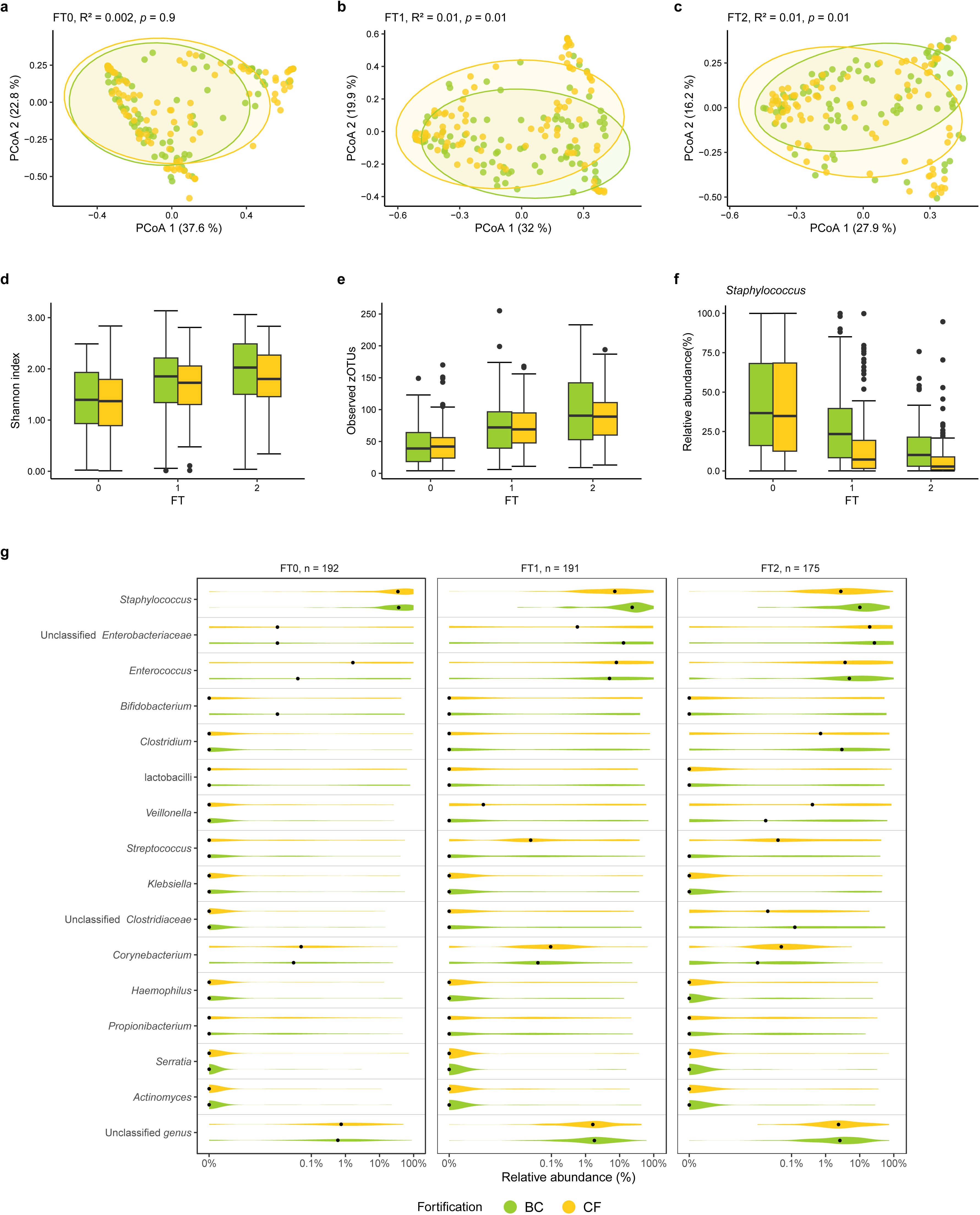
BC or CF as human fortifier has limited effects on the gut microbiota of VPIs. **a-c** The microbial community structure differs between infants receiving BC and CF fortification. A significant difference between the fortifier groups was found at FT1 and FT2, but the variance explained by fortifiers was small. PCoA was based on the weighted Unifrac dissimilarity metrics. R^2^ and *p* values were calculated by PERMANOVA (999 permutations) and adjusted for confounders including GA, SGA, birth mode, DOL, hospital and use of antibiotics. The ellipses in the plots represented 95% CI. **d-e** The species diversities based on the Shannon index and the number of observed zOTUs. No significant difference was found between the BC and CF groups at each FT when compared using linear regression adjusted for the confounders. **f** Relative abundance of *Staphylococcus* in the BC and CF groups. The significant difference in relative abundance as revealed by DESeq2 with adjustment for confounders, disappeared after FDR correction. **g** Relative abundance of the top 15 abundant genera in the BC and CF groups. The order of genera was based on the mean abundance. Median values are shown as black dots. A pseudocount of 1 × 10^-6^ was added to all relative abundance values to facilitate log-scale transform.

The relative abundance of the 15 most abundant genera in the GM of the enrolled preterm infants can be seen from **Fig. 3g**. The relative abundance of *Staphylococcus* was significantly higher in the BC infants, relative to the CF infants, at FT1 (*p* < 0.05) and FT2 (*p* = 0.05, Deseq2), before but not after FDR correction (q > 0.05, FT1 and FT2, **Fig. 3f**).

To assess the influence of base feed (MOM and DHM) on the effect of fortifier (BC or CF) on the GM, 3-day proportion of MOM (n = 202) was included in the analyses as a confounding factor. Fortifier (BC vs. CF) was still found to affect the microbial community structure at FT1 and FT2, with the GM variation explained by fortifier and by the MOM proportion similar (all R^2^ ≈ 0.01, **Table S4**).

### Relative abundance of *Staphylococcus* correlated with bodyweight gain

At FT0 *Staphylococcus* dominated the GM of almost all enrolled VPIs. At FT1 and FT2 *Staphylococcus* was still among the dominant taxa, but showing a decreasing trend in relative abundance (**Fig. 3g**). Interestingly, the change of *Staphylococcus* relative abundance (as determined by the ratio of log10-transformed relative abundance) was negatively correlated with bodyweight gain (delta bodyweight Z-scores) from FT0 to FT2 among all infants (n = 131, q = 0.049, R = -0.28, Pearson correlation, **Fig. 4a**) indicating that weight gain was higher in the VPIs where *Staphylococcus* relative abundance decreased the most from FT0 to FT2. We also tested if similar observations were seen for species diversity and the two other dominating taxa, *Enterococcus* and unclassified *Enterobacteriaceae*, but neither was associated with changes in bodyweight, body length or head circumference (all q > 0.05, Pearson correlation).

**Fig 4.**
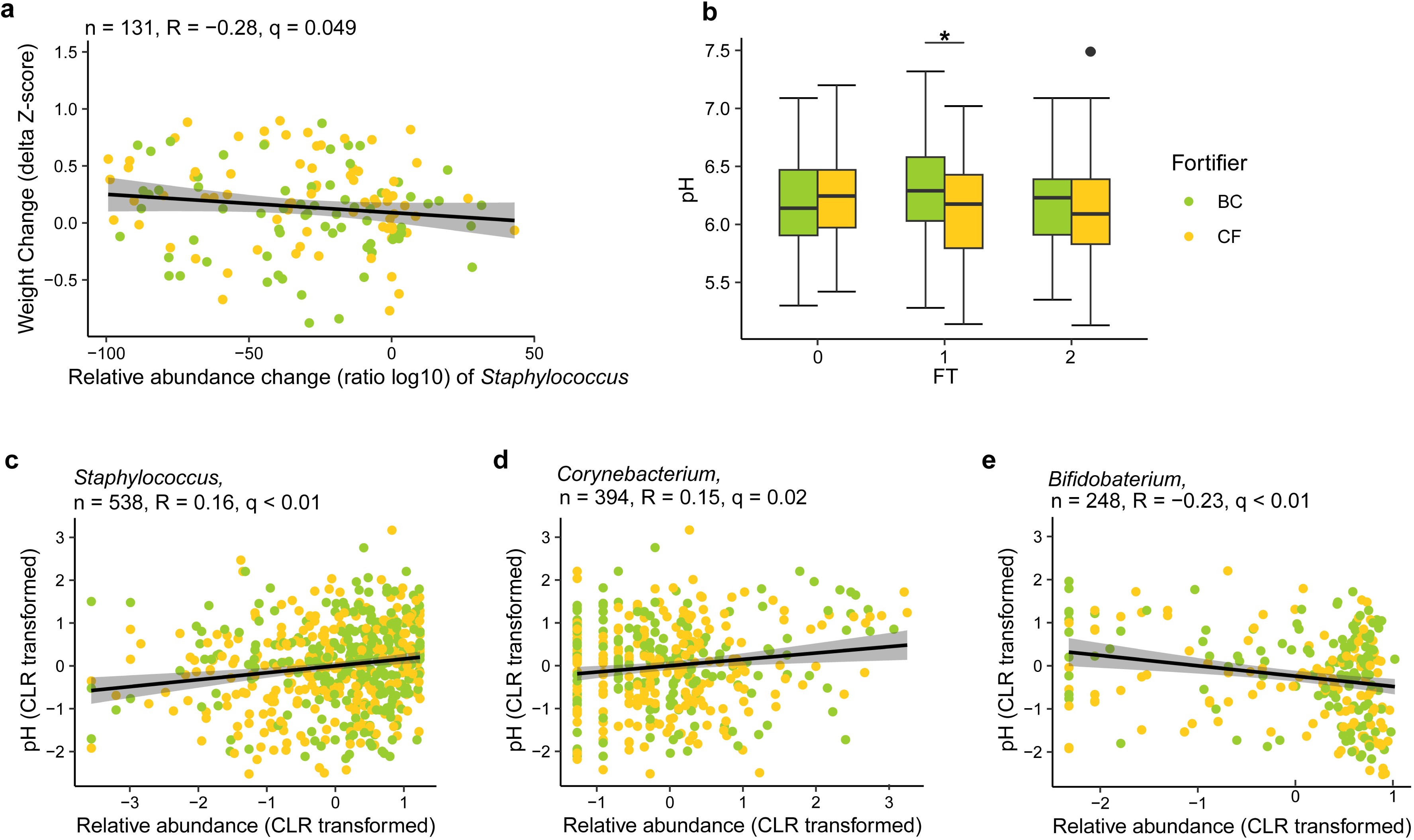
The gut microbiota is correlated with weight gain or fecal pH. **a** The change of *Staphylococcus* relative abundance (as determined by the ratio of log10- transformed relative abundance) was negatively correlated with bodyweight gain (delta bodyweight Z-scores) from FT0 to FT2 across the fortifier groups after FDR correction. Only 131 infants with samples and bodyweight data available at both FTs and within three standard deviations (SDs) were included. **b** Fecal pH was significantly higher in the BC group than in the CF group at FT1 (linear regression). *, *p* < 0.05. **c** Relative abundance of *Staphylococcus* was positively correlated with faecal pH across FTs (q < 0.01, R = 0.2, n = 538). **d** Relative abundance of *Corynebacterium* was positively correlated with faecal pH across FTs (q = 0.02, R = 0.2, n = 394). **e** Relative abundance of *Bifidobacterium* was negatively correlated with faecal pH across three FTs (q < 0.01, R = -0.2, n = 248). Relative abundance of the 15 most abundant genera (central log-ratio transformed) and the pH of faecal samples was correlated by Pearson correlation. *p* values of all correlations were further adjusted by FDR correction to generate q values.

### Choice of fortifier influence fecal pH

Fecal pH was higher in the BC group than in the CF group at FT1 (6.29 ± 0.40 vs 6.14 ± 0.45, mean ± SD, BC vs CF, *p* = 0.02, **Fig. 4b**). The same trend was seen at FT2, though it was not significant. The relative abundance of *Staphylococcus* and *Corynebacterium* was positively correlated with fecal pH (q < 0.01, R = 0.16; q = 0.01, R = 0.15, respectively, Pearson correlation, **Fig. 4c and 3d**), meaning that the higher the fecal pH, the higher the relative abundance of these two genera. The relative abundance of *Bifidobacterium* was on the other hand negatively correlated with fecal pH across fortifier groups and FTs (q < 0.01, R = -0.23, Pearson correlation, **Fig. 4e**).

## Discussion

The first weeks after start of fortification of human milk is a critical time to secure adequate growth of VPIs without overloading the immature gut which could otherwise predispose for intestinal complications like feeding intolerance and NEC [34]. Such complications have been speculated to involve adverse fortification-induced changes to the GM [7,35]. Here we investigated if fortification with two different fortifiers (BC with intact proteins and immunoglobulins, CF with hydrolysed whey protein) influenced short-term GM development and coupled these data with our previous reports on clinical variables and growth outcomes of the same VPIs [19]. The choice of fortifier affected GM composition but the effects were modest and not associated with significant differences in abundance of specific GM genera. In contrast, birth mode (CS or VB) had more pronounced impact, particularly soon after birth, and differences between the two fortification groups were only present among CS infants.

Birth mode is well-known to affect GM in term infants [36,37] while effects in VPIs are more variable [27]. We find that birth mode clearly affected the GM of VPIs shortly after birth but the effect was transient and no longer clear at FT2 at 3-4 weeks of age (**Fig. 2a-c**). The CS VPIs harboured a GM dominated by Gram-positive *Firmicutes* members, while a larger fraction of the VB infants harboured a GM dominated by Gram-negative *Proteobacteria* (**Fig. 2d**). This was also reflected in a tendency for the relative abundance of *Staphylococcus* to be higher in CS versus VB VPIs, especially in the first weeks of life (**Fig. 2e**). It can be speculated that the observed GM differences between CS and VB infants reflect that the CS GM is more dominated by microbes of hospital environment and maternal skin origin, such as *Staphylococcus* [21,38]. The bacteria more frequently found in the GM of term CS infants have been associated with morbidities later in life, such as asthma, infectious and inflammatory diseases, as well as obesity, suggesting that these adverse outcomes observed in CS infants may be attributed to, in part, the GM [39]. However, whether these associations are also evident for preterm infants is uncertain and well-designed studies are necessary to verify the relationship between birth mode-related GM and morbidities. Importantly, our findings suggest that the GM of VPIs born by CS might reconstruct to resemble that of VB infants within the first month of life, potentially alleviating the influence of an aberrant GM after CS on the later disease susceptibility of these infants.

Previous results on the GM effects of fortification in VPIs are conflicting. Two recent studies comparing GM after supplementation with fortifiers based on human or bovine milk, reported significant GM effects [7,35] while another study showed no effect [40]. It is important, however, not only the evaluate effects but also their magnitude. While we found significant BC versus CF effects in our study we conclude that the effect was modest. The variance explained by fortifier type accounted for only ∼1% of the total variance of the microbial community structure, and other factors such as age and weight at birth, birth mode, and antibiotics, could be equally important to GM development than type of fortifier (**Table S3**). Additionally, the difference induced by the fortifiers was primarily observed in the CS infants rather than VB infants, implying that the effects of fortifiers on the GM of the trial may mainly originate from the CS infants.

It has been suggested that the human milk base diet (MOM, DHM or other) has more pronounced effects on shaping the GM in VPIs than milk fortifiers [40]. When we included the 3-day proportion of MOM (as a measure of base feed type) in the statistical models, this did not influence the results. The variance explained by fortifier and base feed in the GM community structure was similar (R^2^ ≈ 0.01), suggesting comparable influence of fortifiers and base feed on the GM composition at least in the time period of our trial. Marked effects of base diet on GM may develop later, consistent with the longer period of observation in the earlier study (DOL 32-60) [40]. Like for fortifiers, GM differences related to base diet may be explained by both the amount and composition of protein. In our study, the VPIs fortified with BC received ∼10% more dietary protein than CF infants [19] and it remains unknown if the differences in GM composition between groups were due to higher protein load or BC-related differences in dietary protein composition [41,42]. In our study, protein derived from fortifiers accounted for 21-22% of the daily protein intake at FT1, and this proportion increased to 26% at FT2 when fortification amount reached a plateau 3-4 weeks after birth. Together the studies suggest that differences in both the human milk base diet and choice of fortifier exert only modest effects on GM composition in the first month of life, the time when the risk of GM-related gut complications is highest.

*Staphylococcus* is considered among the pioneer colonizers of the gut in VPIs, declining in abundance with advancing age and gut maturation [27,43]. The finding that they were found with the highest abundance in infants fortified with BC may reflect a delayed GM development in these infants, potentially induced by slightly higher intake of intact proteins, including immunoglobulins. Interestingly, we found that the relative abundance of *Staphylococcus* was negatively correlated with bodyweight gain, consistent with previous results by Aguilar-Lopez et al. [35]. Yet, a recent meta-analysis could not confirm a consistent link between *Staphylococcus* abundance and bodyweight gain [44], and the significance of these findings for BC-fortified VPIs remains to be clarified.

Fecal pH values reflect changes in the gut environment, potentially derived from diet, GM and/or metabolites [45]. Lower fecal pH is associated with higher colonization resistance against pathogenic bacteria [46], higher abundance of bifidobacteria [47] and bioavailability of minerals, especially calcium [48]. In our study, higher faecal pH in BC-fortified VPIs was associated with higher relative abundance of *Staphylococcus* and lower *Bifidobacterium* abundance. The latter is in accord with previous reports [47,49], possibly reflecting that *Bifidobacterium* are efficient acetate producers. If some intact proteins from BC, such as casein and immunoglobulins, escape digestion in the small intestine, these protein will be excreted or fermented in the colon with the release of alkaline metabolites like ammonia potentially increasing pH [50].

The present study represents the hitherto largest analysis of the effect of a dietary fortification intervention on the GM of VPIs. The samples included were from a well-designed and well-conducted clinical trial with detailed clinical conditions recorded and being available for confounder adjustment in statistical analysis to secure a credible assessment of the effect of fortifiers. Our data are restricted to VPIs in a high-income setting with access to DHM. Lacking information about digestibility of fortifiers complicates our interpretation of the GM changes observed. Another limitation is the amplicon sequencing approach based on the V3 hypervariable region. As for all common amplicon sequencing approaches using short-read technology like the Illumina platforms, accurate species identification is challenging.

## Conclusions

The present study shows that the choice of fortifier, BC or CF, does influence GM composition in VPIs early in life, but also that these effects are to a modest extent. In contrast, the birth mode is a determinant of the GM in this period with a diminishing influence over time. Our finding is in line with other reports and corroborates the notion that the choice of fortifier has limited effects on GM development in the first month of life.

## Methods

### The trial and collection of fecal samples

VPIs included in this study were enrolled in the FortiColos trial. These VPIs were with GA between 26+0 and 30+6 weeks, required nutrient fortification and were from eight neonatal intensive units in Denmark. EN was provided following the local guidelines prior to fortification. When MOM was not available or insufficient, banked DHM was used. Fortification started when EN volume reached 100-140 mL/kg/d and BUN < 5 mmol/L. When BUN was ≥ 5 mmol/L, fortification was delayed or paused until BUN < 5 mmol/L. BC (BC group, ColoDan powder, Biofiber-Damino, Gesten, Denmark) was compared with a conventional BMBF (CF group, PreNan FM85, Nestlé, Switzerland). Initially, 1.0 g of fortifier was added to 100 mL of human milk and gradually increased to a maximum of 2.8 g BC /100 mL and 4.0 g CF /100 ml, both equivalent to the maximum of 1.4 g protein/100 mL (**Fig. 1a**) [19,51]. The fortification continued until the infants reached PMA 34+6 weeks, were early discharged, were moved to a non-participating unit or had diseases. Probiotics (Bifiform, Ferrosan, Denmark), containing freeze-dried LGG (2 × 10^9^ CFU) and BB-12 (2 × 10^8^ CFU), were routinely used in four participating hospitals according to local guidelines (**Table S2**).

Characteristics of enrolled infants including demographics and medical treatment until the end of intervention were prospectively collected from electronic medical records. Stool samples were collected from infant diapers at three time points, including before (FT0), and 1 (FT1) and 2 weeks (FT2) after the fortification (**Fig. 1a, Fig. S1a**). The samples were stored at -50 to -80 °C or placed in refrigerator (+4 °C) for a maximum of 24 hours before being transferred to a -50 or -80 °C freezer.

### Fecal pH measurement

Fecal pH was measured in 1:1 (200 mg:200 mg) diluted samples at room temperature by pH sensor (InLab Micro Pro-ISM equipped with SevenCompact pH meter S220, both from Mettler Toledo, Columbus, USA).

### GM analysis

The fecal GM was profiled by 16S rRNA gene V3 hypervariable region amplicon sequencing. DNA from approximately 200 mg of each sample was extracted by DNeasy PowerSoil Pro Kit (QIAGEN, Hilden, Germany) according to the manufacturer’s instruction. Library preparation was followed the published protocol [52]. The V3 region (Forward primer NXT338: 5’- CCTACGGGWGGCAGCAG-3’, reverse primer NXT518: 5’-ATTACCGCGGCTGCTGG-3’) was used for the two-step PCR amplification. The PCR products were purified with Agencourt AMPure XP Beads (Beckman Coulter Genomics, 245 MA, USA) and quantified with Qubit 1× dsDNA HS assay kit (Invitrogen, Thermo Fisher Scientific). PCR products were pooled in even concentrations and were pair-ended sequenced (2 × 150 bp) by NextSeq (Illumina, San Diego, CA, USA). Sterile DNA-free water was used as negative control and mock community DNA was used as positive control during DNA extraction and library preparation. Both negative and positive controls were included for sequencing.

The bioinformatics process of the sequencing data adhered to established procedures previously described [53]. Specifically, the initial steps involved demultiplexing, merging, and trimming of the raw sequencing data, followed by the removal of chimera and generation of zOTUs using the UNOISE3 [54] algorithm implemented in Vsearch (version 2.21.1) [55]. The Greengenes (13.8) [56] 16S rRNA gene database was employed as a reference database for taxonomic annotation. The mean and median sequencing depth of all samples were 91,251 and 82,865 reads, respectively. In total, 2,535 unique zOTUs were identified in all samples with 2,227 in the BC group and 2,351 in the CF group, respectively.

### Data analysis

Characteristics of included infants including continuous and categorical variables were described by mean (standard deviation) and count (percentage), respectively. Group comparisons were conducted using one-way analysis of variance (ANOVA) for continuous variables and chi-square (χ^2^) test for categorical variables.

Analysis and visualization of the microbiota data were performed by R packages Phyloseq [57], Vegan [58] and ggplot2 [59]. The raw zOTUs were rarefied at an even depth of 10,000 counts per sample. The samples with insufficient sequencing depth were excluded from the analysis (5 samples, **Fig. 1b**).

The species diversity (alpha-diversity), measured by the number of observed zOTUs, was compared between the birth mode groups (CS vs. VB) by linear regression with adjustment for confounders, including fortifier, GA, SGA, DOL, hospital and use of antibiotics. The microbial community structure (beta-diversity) was assessed based on weighted UniFrac distance dissimilarity metrics and shown with principal co-ordinates analysis (PCoA) score plot. The group difference (CS vs. VB) was tested by permutational multivariate analysis of variance (PERMANOVA). All samples were classified into three GM types according to the dominating phylum with relative abundance over 50%: *Firmicutes*-dominated, *Proteobacteria*-dominated and a other pattern. Each GM type of the CS and VB groups at the same FT was compared by chi-square (χ^2^) test. The comparison of infants as *Firmicutes*-dominated or not between different FTs was conducted using logistic regression with adjustments for confounders within each birth mode group. DESeq2 with the same covariates was used to locate OTUs with different relative abundance between two groups[60] (only tested if relative abundance was over 1% and present in over 50% of samples in either fortifier group at each FT). FDR approach was used to correct *p* values from multiple tests at each FT.

Comparison between the fortifier groups (BC vs. CF) on the species diversity assessed by the Shannon diversity index and the number of observed zOTUs), microbial community structure assessed based on weighted UniFrac, unweighted UniFrac, Bray Curtis and Jaccard (dis)similarity metrics, and OTUs with different relative abundance was conducted by the same methods mentioned above with adjustment for different confounders, including birth mode, GA, SGA, DOL, hospital and use of antibiotics. A parameter, 3-day proportion of MOM, was defined as the number of meals of exclusive MOM over the number of all meals in the three days before a FT. A meal with both MOM and DHM is registered as 50% MOM. This parameter was included in the analyses mentioned above to adjust for the effect of base feed on the GM.

Changes in relative abundance of the three most abundant bacterial genera (*Staphylococcus, Enterococcus* and unclassified *Enterobacteriaceae*) and species diversity (the Shannon index and the number of zOTUs) were correlated with anthropometric parameters (bodyweight, body length and head circumference) between different FTs across the fortifier groups. Bodyweight, body length and head circumference was calculated into Z-scores with reference to the Swedish growth charts for preterm infants [61]. Relative abundance was log10-transformed. Changes were assessed by delta for anthropometric parameters and species diversity, or ratio for the relative abundance between two FTs, respectively. Pearson correlations were adapted after removing outliners of ratio according to three-sigma limits and FDR approach was used to correct *p* values within the same period. Relative abundance of the 15 most abundant genera was correlated with the pH of fecal samples using Pearson correlation with central log-ratio (CLR) transformation [62]. FDR approach for the correlation analysis between relative abundance and pH was adapted for *p* values supported by at least 30% of samples.

All statistical analyses were performed in R (version 4.2.1). FDR-corrected *p* values were shown as q values. The *p* or q < 0.05 was regarded statistically significant.

## Data Availability

The 16S sequencing data produced in the present study is available in the Sequence Read Archive at NCBI with the accession number PRJNA1090537 in an anonymized form.
Clinical data containing personally identifiable information of individual infants participating in the cohort cannot be freely available to protect the privacy of the participants and their families. This is in accordance with the Danish Data Protection Act and European Regulation 2016/679 of the European Parliament and of the Council (GDPR), which prohibit distribution even in pseudo-anonymized form.

## Declarations

### Ethics approval and consent to participate

The FortiColos study received approval from the Scientific Ethical Committee of the Region of Southern Denmark (S-20170095) and the Danish Data Protection Agency (17/33672). An independent data safety monitoring board (DSMB) followed and reviewed trial data and safety. Parental consent was obtained in writing from both parents of participated infants prior to intervention.

### Availability of data and materials

The 16S sequencing data supporting the conclusions of this article is available in the Sequence Read Archive at NCBI with the accession number PRJNA1090537 in an anonymized form.

Clinical data containing personally identifiable information of individual infants participating in the cohort cannot be freely available to protect the privacy of the participants and their families. This is in accordance with the Danish Data Protection Act and European Regulation 2016/679 of the European Parliament and of the Council (GDPR), which prohibit distribution even in pseudo-anonymized form. However, research collaborations are welcome, and access to the data can be made through a joint research collaboration by contacting Gitte Zachariassen.

All code to analysis data and generate the presented results and manuscript figures is available via https://github.com/linyang1106/FortiColos_microbiota

### Authors’ Contributions

P.T.S, G.Z. and L.A. contributed to the design. G.Z. and L.A. contributed to the conduction of the trial and the acquisition of samples and data. L.Y. and Y.H. contributed to the lab experiment. W.P.K. contributed to the sequencing. L.Y., Y.H., P.P.J. and D.S.N. contributed to the analysis and visualization. L.Y. and P.P. contributed to the draft of the manuscript.

L.Y., Y.H., P.P.J., D.S.N., P.T.S., L.A. and G.Z. contributed to the interpretation and revision. All authors have read and agreed to the final version of the manuscript.

## Acknowledgements

We express our great gratitude to all the infants and families for their contributions. We acknowledge AM Ahhnfeldt, SS Kappel and all the participating units and clinicians for their supports. We acknowledge DV Stefanova for laboratory supports.

