## Supplementary material for "Gut microbiota development in very preterm infants following fortification of human milk": Fig. S1; Fig. S2; Fig. S3; Fig. S4; Fig. S5; Table S1; Table S2; Table S3; Table S4.

### Supplementary Figures

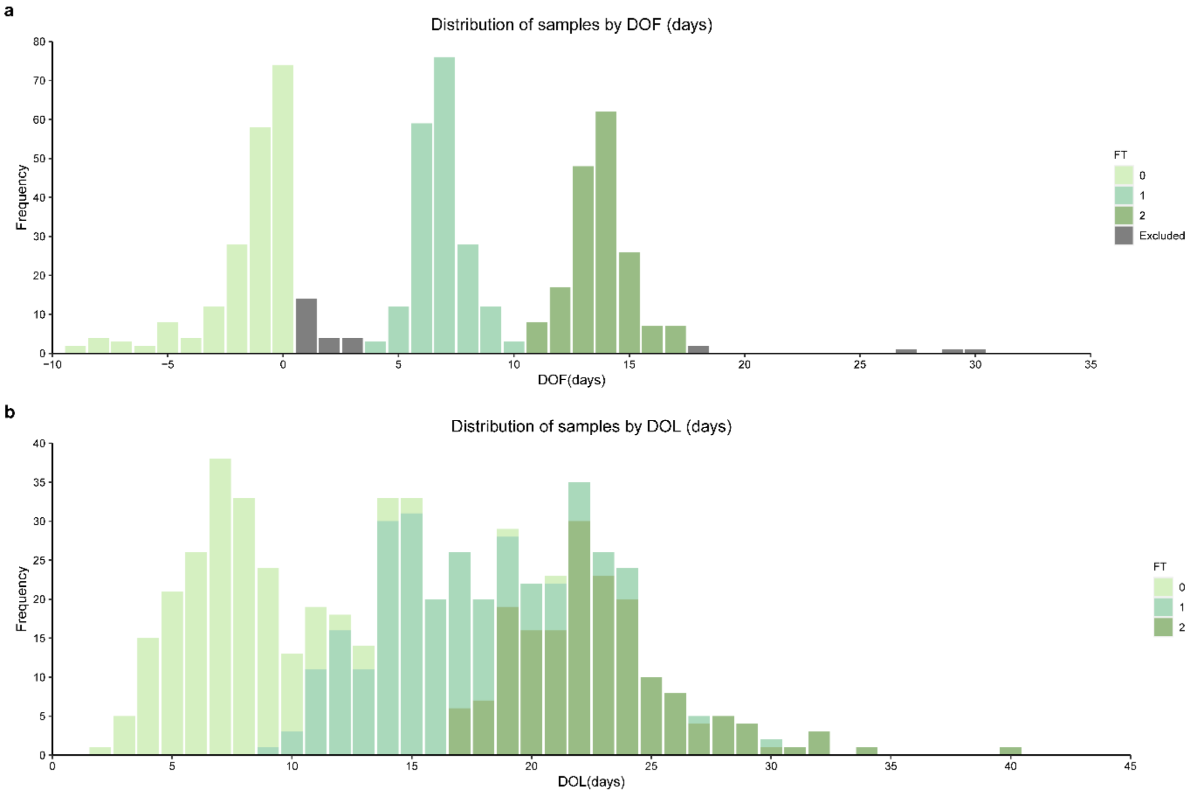


**Fig. S1: Distribution of samples by days of fortification (DOF) or days of life (DOL).**

**a** The distribution of samples according to DOF. Samples were included at three time points: before fortification (from -10 to 0 days of fortification, FT0), around one week (7 ± 3 DOF, FT1) and two weeks of fortification (14 ± 3 DOF, FT2). **b** The distribution of samples according to DOL. The DOL of samples included in FT0, FT1 and FT2 roughly corresponded to 2-10, 10-20 and 20-30 days, respectively.

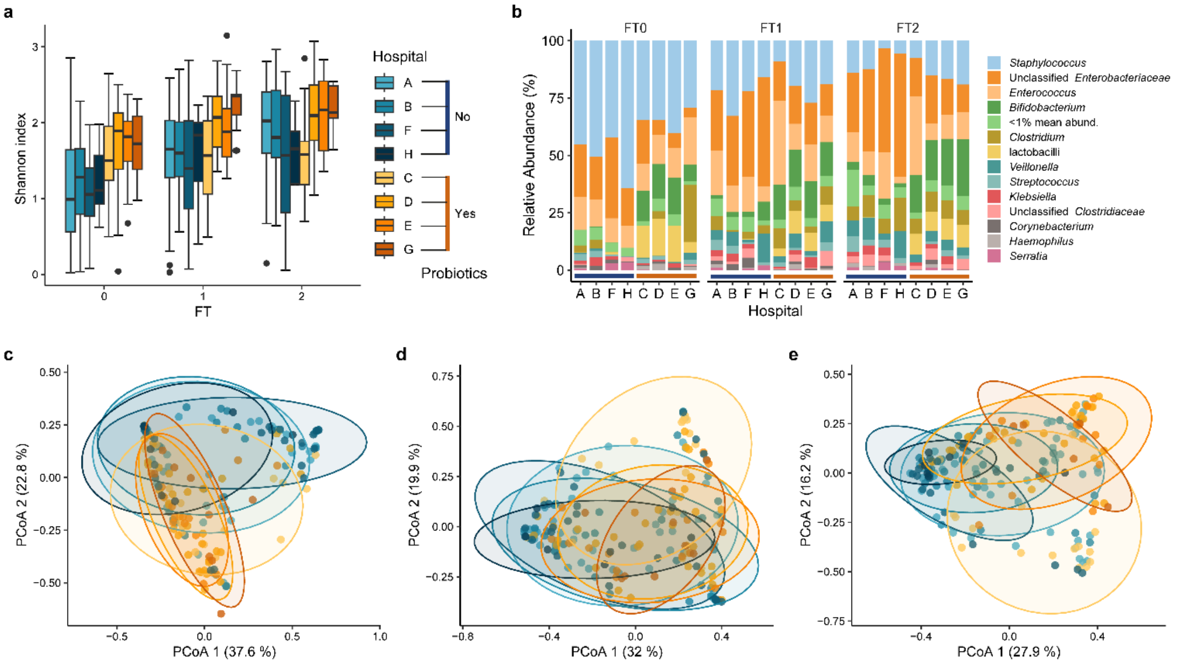


**Fig. S2 Hospitals intricately alter the gut microbiota with probiotics.**

**a** Species diversity was assessed by the Shannon index in different hospitals. **b** The relative abundance of different genera according to hospital groups. Taxa that could not be classified as a specific genus were labelled "unclassified" with their upper-level taxa. Genera with mean relative abundance lower than 1% at each FT was summed up and labelled as "<1% mean abund.". **c-e** PCoA plot representing microbial community structure based on weighted Unifrac distance of different hospital groups. The ellipses represented 95% confidence intervals.

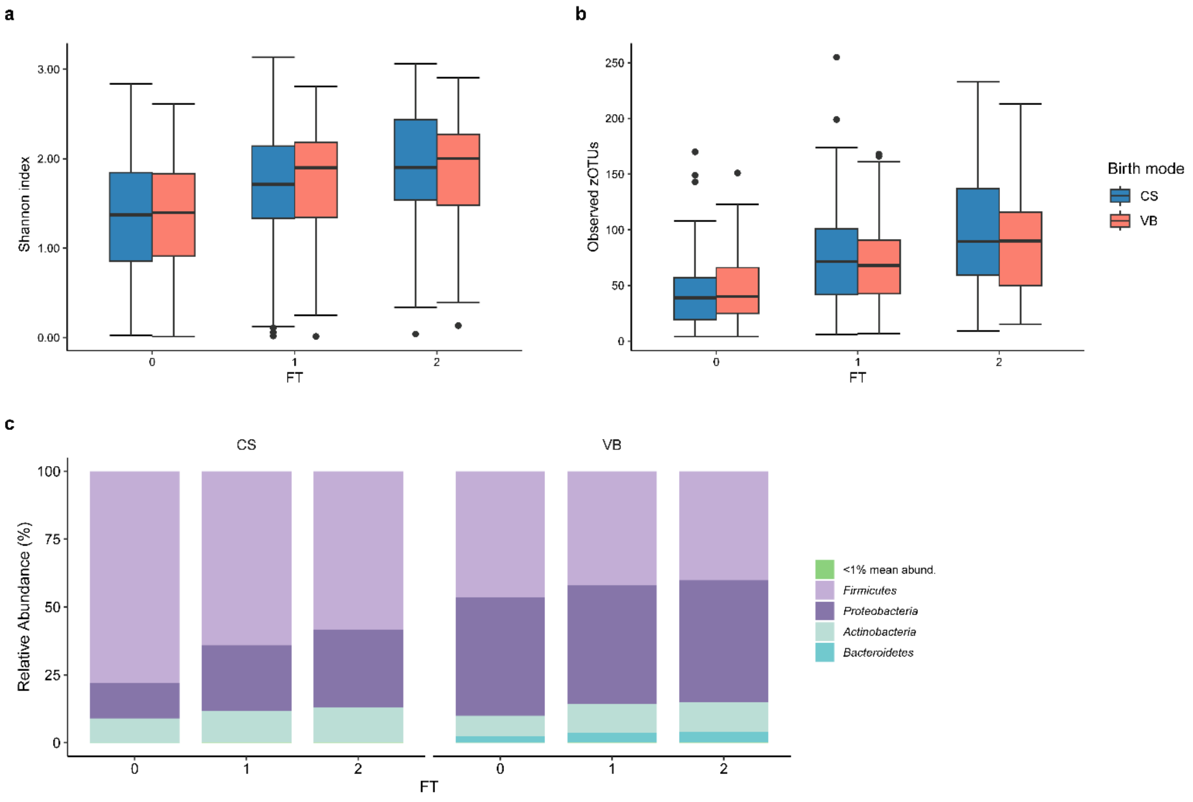


**Fig. S3 Species diversity and the relative abundance at phylum level in VPIs born by CS and VB.**

**a-b** No significant difference in species diversity between VB and CS groups. The species diversity based on the Shannon index at different FTs was compared by linear regression adjusted for confounders including GA, SGA, fortification, DOL, hospital and use of antibiotics. **c** The relative abundance of different phyla of different birth mode groups. *Bacteroidetes* were predominantly identified in infants delivered vaginally. Phyla with mean relative abundance lower than 1% at each FT was summed up and labelled as "<1% mean abund.".

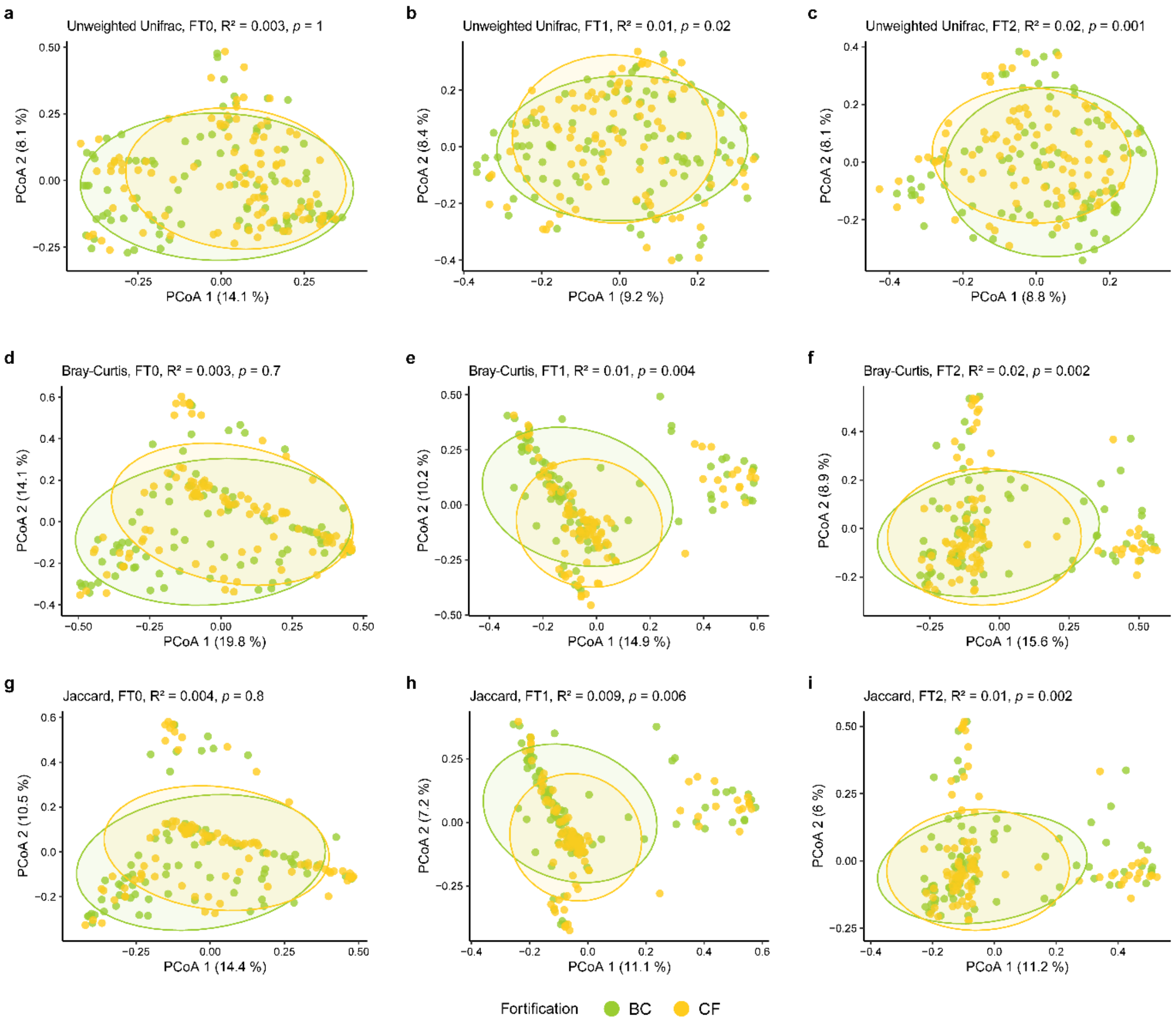


**Fig. S4 Microbial community structure assessed by different (dis)similarity metrics comparing BC and CF infants.**

**a-c** PCoA figures based on unweighted Unifrac distance. **d-f** PCoA figures based on Bray Curtis distance. **g-i** PCoA figures based on Jaccard distance. Significant difference was found after fortification started between BC and CF groups. The ellipses represented 95% confidence intervals. $R^2$ and *p*-values were calculated between BC and CF groups by PERMANOVA (999 permutations) and adjusted for confounding factors including GA, SGA, birth mode, DOL, hospital and use of antibiotics.

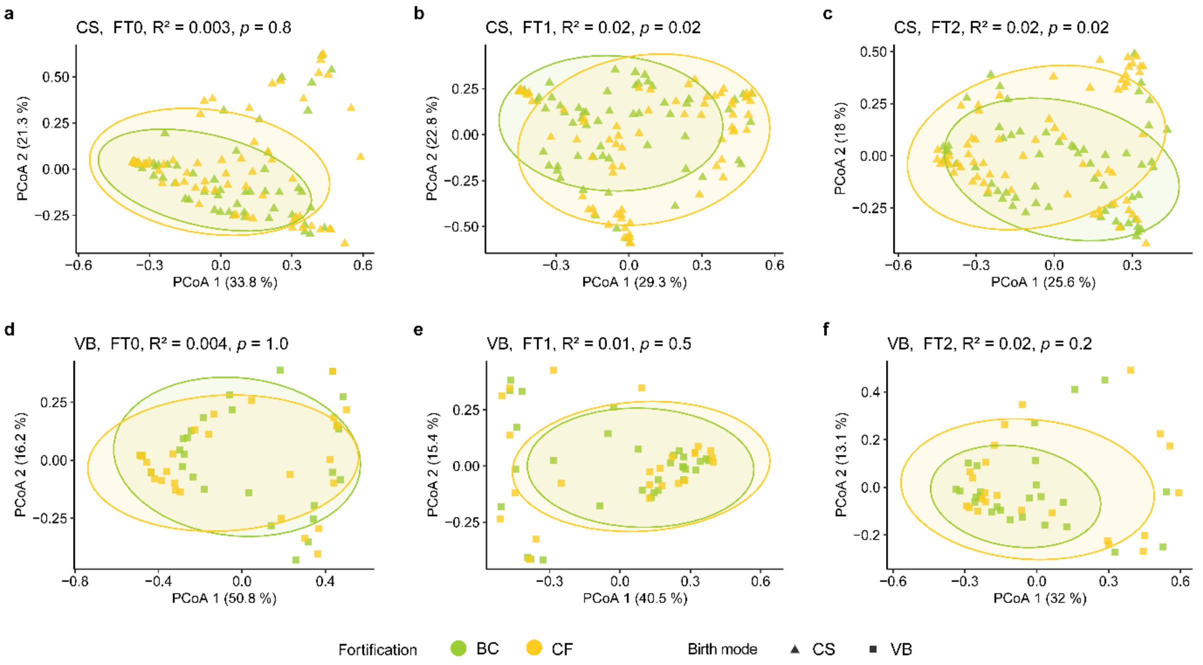


**Fig. S5 The choice of fortifier only influenced the GM of CS infants but not VB infants.**

**a-c** PCoA plot presenting microbial community structure of BC and CF groups in CS infants. Microbial community structure was significantly different between fortifier groups in CS infants. **d-f** PCoA plot presenting microbial community of BC and CF groups in VB infants. No significant difference was found between groups in VB infants. PCoA was based on weighted Unifrac distance. The ellipses represented 95% confidence intervals. $R^2$ and *p* values were calculated by PERMANOVA (999 permutations) and adjusted for confounders including GA, SGA, DOL, hospital and use of antibiotics.

**Supplementary tables**

**Table S1. Characteristics of infants based on birth mode**

| **Characteristics** | **CS** | **VB** | ***p* value** |
|---|---|---|---|
| N | 161 | 64 | |
| GA, weeks (mean (SD)) | 28.73 (1.46) | 28.54 (1.44) | 0.386 |
| Gender, Male/Female (%) | 92 (57.1)/69 (42.9) | 39 (60.9)/25 (39.1) | 0.711 |
| Birth weight, g (mean (SD)) | 1139.90 (328.68) | 1250.80 (309.50) | 0.021 |
| SGA[1], Yes/No (%) | 49 (30.4)/112 (69.6) | 1 (1.6)/63 (98.4) | <0.001 |
| Multiple birth, Yes/No (%) | 50 (31.1)/111 (68.9) | 19 (29.7)/ 45(70.3) | 0.968 |
| Fortification, BC/CF (%) | 74 (46.0)/87 (54.0) | 35 (54.7)/ 29(45.3) | 0.301 |
| Probiotics use[2], Yes/No (%) | 48 (29.8)/113 (70.2) | 116(25.0)/48 (75.0) | 0.577 |
| 3-day MOM proportion, % (mean (SD))[3] | | | |
| FT 1 | 84.54 (28.10) | 88.53 (23.52) | 0.368 |
| FT 2 | 83.83 (30.94) | 92.16 (21.09) | 0.088 |
| Antibiotics use[4], Yes/No (%) | | | |
| FT 0 | 61 (43.9)/78 (56.1) | 16 (30.2)/37 (69.8) | 0.117 |
| FT 1 | 30 (22.1)/106 (77.9) | 8 (14.5)/47 (85.5) | 0.328 |
| FT 2 | 17 (13.5)/109 (86.5) | 7 (14.3)/42 (85.7) | 1.000 |

1. SGA is defined as a birth weight (BW) Z score ≤ -2 standard deviations for GA.
2. Probiotics use is defined as using probiotics before intervention started.
3. Feeding data was recorded after fortification start.
4. Antibiotics use is defined as using antibiotics within 5 days prior stool sample collated.

**Table S2 The use of probiotics before fortification in different hospitals**

| Hospital | Yes (%) | No (%) | Total |
|---|---|---|---|
| A | 0 (0.0) | 58 (100.0) | 58 |
| B | 0 (0.0) | 32 (100.0) | 32 |
| C | 11 (44.0) | 14 (56.0) | 25 |
| D | 30 (69.8) | 13 (30.2) | 43 |
| E | 14 (66.7) | 7 (33.3) | 21 |
| F | 1 (3.7)* | 26 (96.3) | 27 |
| G | 8 (66.7) | 4 (33.3) | 12 |
| H | 0 (0.0) | 7 (100.0) | 7 |
| Total | 64 | 161 | 225 |

* One infant used probiotics in hospital E and was transferred to hospital F.

**Table S3 Distance-based redundancy analysis of included variables**

| variable | FT0 | | | FT1 | | | FT2 | | |
|---|---|---|---|---|---|---|---|---|---|
| | Sum of squares | F value | *p* value | Sum of squares | F value | *p* value | Sum of squares | F value | *p* value |
| Fortifier | 1.38 | 1.26 | * | 1.46 | 1.65 | *** | 1.77 | 2.13 | *** |
| GA | 1.79 | 1.64 | ** | 1.92 | 2.17 | *** | 1.79 | 2.16 | *** |
| SGA[1] | 1.62 | 1.48 | ** | 1.40 | 1.15 | ** | 1.16 | 1.25 | * |
| Birth mode | 2.28 | 2.08 | *** | 1.30 | 1.42 | * | 1.06 | 1.28 | # |
| DOL | 1.16 | 1.06 | - | 0.96 | 1.08 | - | 1.27 | 1.57 | ** |
| Hospital | 12.8 | 1.67 | *** | 11.3 | 1.83 | *** | 11.2 | 1.94 | *** |
| Antibiotics[2] | 1.92 | 1.75 | *** | 1.56 | 1.77 | ** | 1.14 | 1.35 | * |
| residual | 194.64 | | | 156.75 | | | 133.77 | | |

-, $p \geq 0.1$; #, $p < 0.1$; *, $p < 0.05$; **, $p < 0.01$; ***, $p < 0.001$.

1. SGA is defined as a birth weight (BW) Z score ≤ -2 standard deviations for GA.
2. Antibiotics use is defined as using antibiotics within 5 days prior stool sample collated.

**Table S4 Effects of base feed on microbial community structure**

| Weighted Unifra distance | FT 1 | | FT 2 | |
|---|---|---|---|---|
| | *p* | $R^2$ | *p* | $R^2$ |
| Fortification | 0.01 | 0.01 | 0.02 | 0.01 |
| 3-day MOM proportion | 0.4 | 0.005 | 0.02 | 0.01 |
